# Host response-based screening to identify undiagnosed cases of COVID-19 and expand testing capacity

**DOI:** 10.1101/2020.06.04.20109306

**Authors:** Nagarjuna R. Cheemarla, Anderson F. Brito, Joseph R. Fauver, Tara Alpert, Chantal B.F. Vogels, Saad B. Omer, Albert I. Ko, Nathan D. Grubaugh, Marie L. Landry, Ellen F. Foxman

## Abstract

The COVID-19 pandemic has created unprecedented challenges in diagnostic testing. At the beginning of the epidemic, a confluence of factors resulted in delayed deployment of PCR-based diagnostic tests, resulting in lack of testing of individuals with symptoms of the disease. Although these tests are now more widely available, it is estimated that a three- to ten-fold increase in testing capacity will be required to ensure adequate surveillance as communities reopen^1^. In response to these challenges, we evaluated potential roles of host response-based screening in the diagnosis of COVID-19. Previous work from our group showed that the nasopharyngeal (NP) level of CXCL10, a protein produced as part of the host response to viral infection, is a sensitive predictor of respiratory virus infection across a wide spectrum of viruses^2^. Here, we show that NP CXCL10 is elevated during SARS-CoV-2 infection and use a CXCL10-based screening strategy to identify four undiagnosed cases of COVID-19 in Connecticut in early March. In a second set of samples tested at the Yale New Haven Hospital, we show that NP CXCL10 had excellent performance as a rule-out test (NPV 0.99, 95% C.I. 0.985-0.997). Our results demonstrate how biomarker-based screening could be used to leverage existing PCR testing capacity to rapidly enable widespread testing for COVID-19.

**One Sentence Summary:** We describe a host-response based screening strategy to identify undiagnosed cases of COVID-19 and to expand capacity for PCR-based testing.

## Main

The COVID19 pandemic has disrupted normal activity around the globe and has created unprecedented challenges for diagnostic testing. At the start of the pandemic, establishing testing to detect the virus was a challenge, and currently the challenge is capacity. Many experts estimate a need to expand testing capacity up to ten-fold to ensure adequate surveillance as businesses reopen^1^. While PCR-based detection of viral genomes is the current gold standard for diagnosis of SARS-CoV2 and is highly specific, this method requires trained laboratory personnel, has multiple steps, and uses costly reagents subject to supply chain disruption. Based on previous work, we hypothesized that an elevated level of the interferon inducible protein CXCL10 in the nasopharynx could serve as a sensitive screen for patients with an active respiratory virus infection including SARS-CoV2^2^. If so, this test could be used as a pre-screen to direct PCR testing to patients most likely to be SARS-CoV2 positive, allowing accurate diagnosis while preserving PCR testing capacity. Protein measurements lend themselves to rapid, high throughput, and point-of care diagnostic methodologies^3,4^, so this combined strategy offers the potential to have a first step which is sensitive and convenient (CXCL10 assay) followed by a more specific test (PCR) for a subset of screen-positive patients.

First, we sought to use this strategy to screen for cases of COVID-19 in the Yale-New Haven Healthcare System (YNNHS) beginning in early March of 2020, prior to the start of testing at our institution on March 13 (Fig 1a). The first reported cases of COVID-19 in the region occurred in New York state on March 2nd and in Connecticut on March 9th (Fig S1)^5-7^. We focused on the time window from March 3rd to 14th, 2020. During this time frame, 642 nasopharyngeal (NP) swab specimens were tested by the Yale-New Haven Hospital (YNHH) clinical virology laboratory using a laboratory developed respiratory virus PCR panel (RVP) that detects 15 common respiratory viruses (Table S1). Of 642 NP samples, 376 were negative for all viruses on the panel, but the SARS-CoV-2 status was unknown.

Previously, we found that the CXCL10 level in the NP swab viral transport medium was a useful indicator of infection with many common respiratory viruses^2^, suggesting that this measurement could be used to indicate which of these 376 samples were most likely to contain a virus not detected by the RVP. NP CXCL10 is expected to be undetectable in healthy subjects. First, to assess whether CXCL10 could be used as a biomarker for SARS-CoV-2 infection, we measured CXCL10 concentration in 39 confirmed SARS-CoV-2 positive NP swab specimens from the third week of March and compared to levels in the RVP-negative samples from March 3rd to 14th. SARS-CoV-2 positive samples showed an average CXCL10 concentration of 681pg/ml (Fig 1b). Notably, the two SARS-CoV-2 positive samples with lowest CXCL10 levels also had very low virus RNA concentrations by RT-qPCR (Ct values for N1 were 36.4 and 34.9). In contrast, the majority of RVP-negative samples had CXCL10 levels at or below the limit of detection, with only 8.5% (32/376) of the samples having CXCL10 levels above a threshold value of 100 pg/ml (Fig 1b). We hypothesized that some of these samples might be from patients with undiagnosed COVID-19.

**Fig 1.**
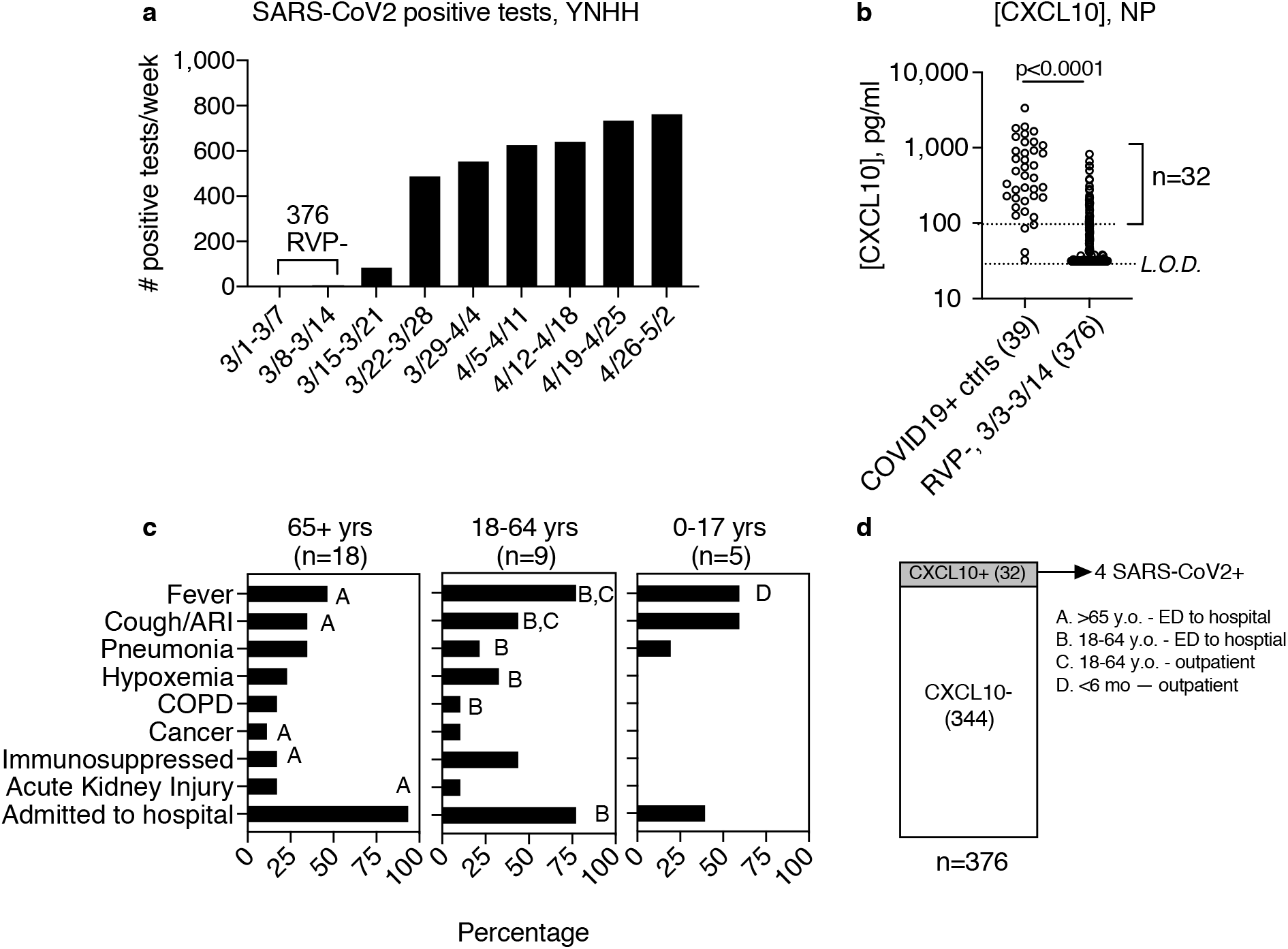
Screen for undiagnosed cases of SARS-CoV-2, March 3-14, 2020. **(a)** Number of positive tests for SARS-CoV-2 by week, Yale-New Haven Hospital (YNHH). Bracket indicates time frame chosen for study, March 3-March 14, during which 376 samples tested negative for viruses on the YNHH 15 respiratory virus panel (RVP). **(b)** CXCL10 concentration was measured in viral transport medium associated with 39 COVID-19+ positive cases from the third week of March excluding March 20 (left) and 376 RVP negative NP swabs from March 3-14 (right). Bracket indicates RVP-samples with CXCL10 concentrations equal to or above the cutoff of 100 pg/ml. Limit of detection (L.O.D) of ELISA assay is shown**. (c)** Clinical presentation of 32 patients from March 3-March 14 testing RVP-, CXCL10+, by age group. Bar indicates percentage of patients in each age category displaying the clinical features listed on the y-axis. Letters A-D indicate clinical features of individual patients who ultimately tested positive for SARS-COV-2 by RT-qPCR. Clinical data was not available for patient B. **(D)** All 376 RVP-samples were screened for SARS-CoV-2 using RT-qPCR as described in the material and methods. SARS-CoV-2 viral RNA was detected in 4 out of 32 CXCL10 positive samples and none of the 344 CXCL10 negative samples, as shown. Patient age, sample collection date, and site of patient care is indicated for each of the four positive samples.

Next, we examined the medical records for patient clinical features associated with the RVP-negative, CXCL10-positive samples (Fig 1c). The majority of samples (18/32) were from patients 65 years of age or older. These patients displayed typical symptoms of COVID-19 and other respiratory infections with the most common being fever, cough, and evidence of pneumonia and/or hypoxemia. Most patients in this age group were admitted to the hospital. Nine samples were from patients ages 18-64 and five were from children, who also typically displayed fever and cough, and to a lesser extent other symptoms, and fewer were hospitalized. Then we tested RNA extracted from CXCL10-positive samples for SARS-CoV-2 RNA using RT-qPCR. Four of the 32 samples contained SARS-CoV-2 RNA, and this result was confirmed by the YNHH clinical virology laboratory (Fig 1d). To evaluate the performance of CXCL10 as a pre-screen for virus-positive samples, we also tested the 344 CXCL10-low samples for SARS-CoV-2 RNA, and all were negative. For completeness, we also tested the 266 RVP-positive samples from this time period for SARS-CoV-2 RNA and all were negative. Thus, the four samples identified in the screen were the only SARS-CoV-2-positive among the 642 NP samples tested by RVP in this time frame, and are among the first SARS-CoV-2-positive samples in our health care system.

The identified SARS-CoV-2-positive samples were collected on March 10^th^-14th. Three of these samples were from adults, two of whom were subsequently hospitalized for COVID-19. Surprisingly, one sample was from a young child seen as an outpatient. Clinical features associated with each patient presentation are indicated by the letters A-D on Figure 1c.

To gain further insight into the epidemiology of these early cases of SARS-CoV-2, we performed whole-genome sequencing and phylogenetic analysis (Fig. 2; data can be visualized at: https://nextstrain.org/community/grubaughlab/CT-SARS-CoV-2/paper3). The full list of genomes included in this analysis and details of genome coverage can be found in Table S2. Interestingly, the four SARS-CoV-2 isolates were genetically distinct. Previous work by our group described a cluster of early cases in Connecticut closely related to strains from Washington State (lineage A1), including the SARS-CoV-2 sequenced from one of these patients (genome Yale-009)^8^. The other newly sequenced positive cases from this screen (genomes Yale-011, 040 and 151) belong to a distinct lineage (B1). Yale-040 groups within the sub-lineage B1.11, which has Western Europe, likely the United Kingdom, as the most probable origin. Finally, the genomes Yale-040 and 011 are closely related to isolates from New York state but group in different clades (Fig. 2), indicating that SARS-CoV-2 entered Connecticut via multiple independent lines of transmission by early March.

**Figure 2.**
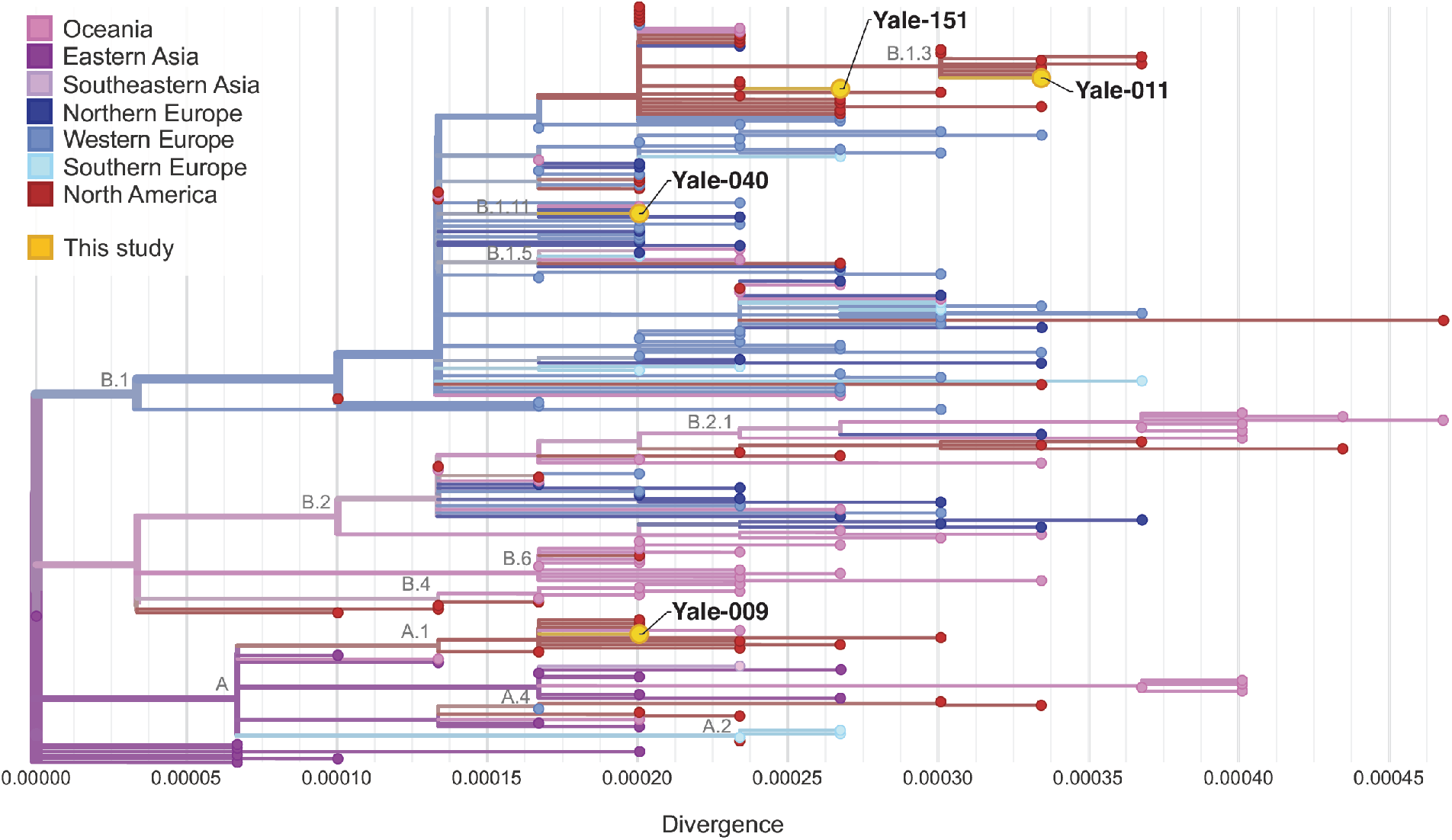
Phylogenetic analysis of four genomes from early March cases of COVID-19 identified with CXCL10-based screening. >Maximum-likelihood phylogenetic tree of the four SARS-CoV-2 genomes sequenced in this study with other 210 genomes obtained from around the world. Further information can be found online at: https://nextstrain.org/community/grubaughlab/CT-SARS-CoV-2/paper3. Acknowledgements of authors of the genomes used in this study can be found in Table S3.

The fact that all four SARS-CoV-2-positive samples were among the CXCL10-high group (Fig 1d) suggested that a CXCL10-based screening strategy might be an efficient way to leverage existing PCR tests to greatly expand testing capacity, due to the high negative predictive value of a negative NP CXCL10 screen. In other words, we would have found all SARS-CoV-2-positive samples even if we had not performed PCR testing on the >90% of samples that were below the 100pg/ml CXCL10 cutoff. To further evaluate the performance of this strategy (biomarker based screen followed by PCR test), we next tested a second group of samples – all samples sent to the YNHH laboratory for COVID-19 testing on a single day in March. We chose March 20th, 2020, since we were able to obtain residual samples for all 144 NP swab samples for which SARS-CoV-2 PCR testing was ordered on that date. Since these samples were not pre-filtered to exclude patients with other viral infections, we expected to see a higher rate of CXCL10-positive samples in this sample set than in the RVP-negative samples. We measured CXCL10 concentration in all samples. Using the cut-off of 100 pg/ml, 35/144 samples were CXCL10-positive and these included 16/17 SARS-CoV-2-positive samples; conversely, 109/144 samples were CXCL10-negative and these included one SARS-CoV-2-positive sample. Notably, this sample had the lowest viral RNA level of all samples tested that day. Examining the relationship between viral load and NP CXCL10 across all 17 SARS-CoV-2-positive samples revealed a significant positive correlation (Fig 3c). Considering all 144 samples evaluated for suspected COVID-19 on March 20, a negative test for CXCL10 had a very high negative predictive value for SARS-CoV-2 infection (NPV=0.99, 95% C.I. 0.985-0.997), indicating its usefulness as a rule-out test.

**Fig 3.**
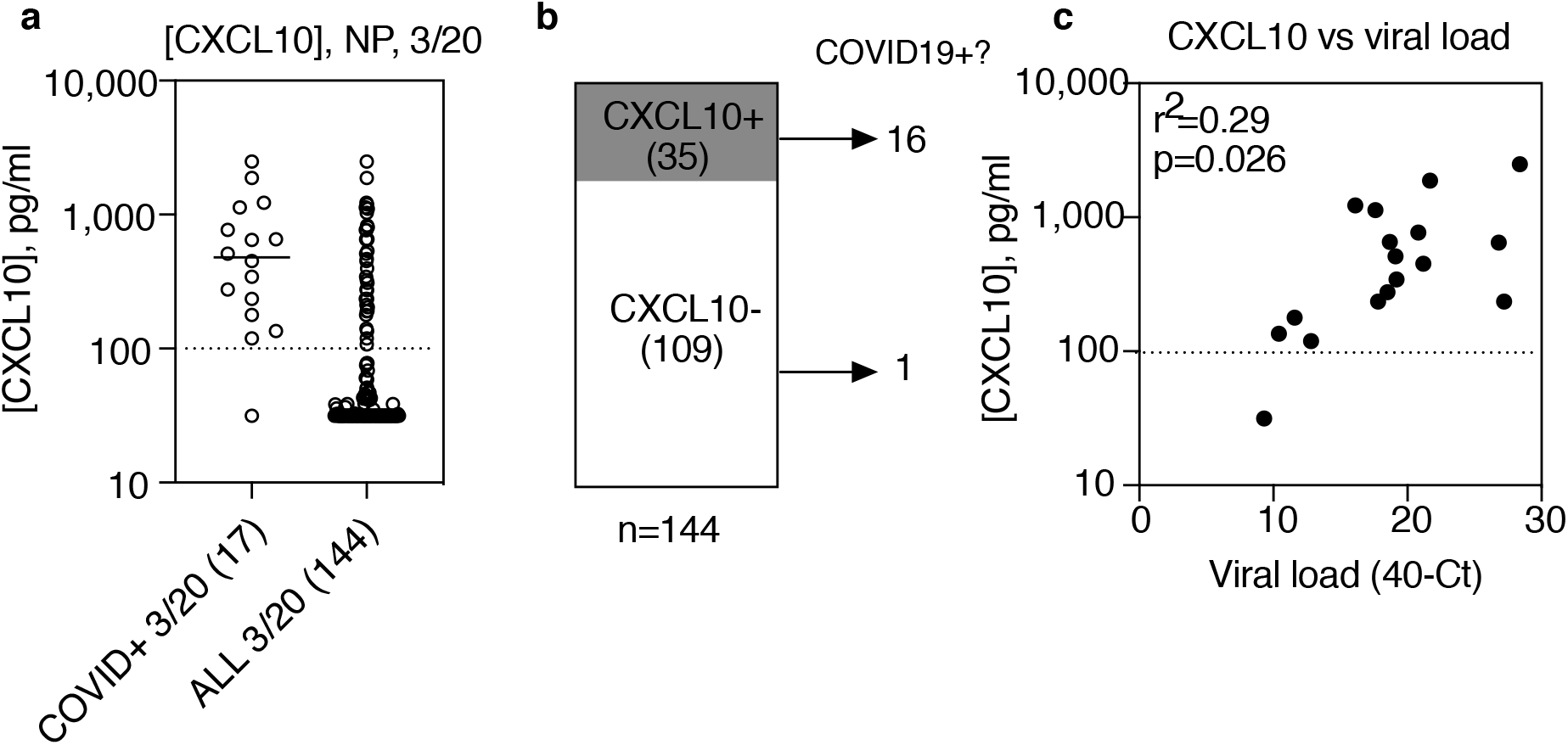
CXCL10 pre-screen in samples sent for SARS-CoV-2 PCR test on March 20, 2020 (n=144) **(a)** CXCL10 concentration was measured in viral transport medium in all 144 samples sent for SARS-CoV-2 PCR testing on March 20, 2020. Scatter plots show [CXCL10] for 17 positive samples only (left) and all samples (right). Limit of detection (L.O.D.) of ELISA assay is 31.5 pg/ml**. (b)** Stacked bars depict proportion of samples above the CXCL10 cutoff of 100 pg/ml and distribution of CXCL10 positive samples. **(c)** Relationship between viral load, expressed at 40-Ct value for N1 gene, and NP [CXCL10] in 17 positive samples. Two-tailed Pearson correlation analysis indicates significant positive association between viral load and NP [CXCL10].

Finally, to better understand the possible uses of CXCL10 in the diagnosis of COVID-19, we studied the correlation between NP CXCL10 level and biological variables using all 59 of the SARS-CoV-2-positive samples in this study (4 from March 3rd to 14th, 17 diagnosed on March 20th, and 39 positive control from other dates during the third week of March) (Fig 4a, b). CXCL10 is a chemokine involved in attracting T cells to sites of viral infection, and is a classic interferon stimulated gene (ISG), a gene highly induced by interferon signaling or by activation of intracellular sensors for viral RNA within infected cells^9,10^. Recent studies of innate immune response to SARS-CoV-2 indicate that the virus triggers a robust antiviral interferon response in the airway^11-14^. Therefore, we reasoned that NP CXCL10 level might reflect the level of active viral replication, the stimulus for CXCL10 production. Consistently, across all SARS-CoV-2+ samples in the study, viral load was positively associated with NP CXCL10 (Fig 4c).

**Fig 4.**
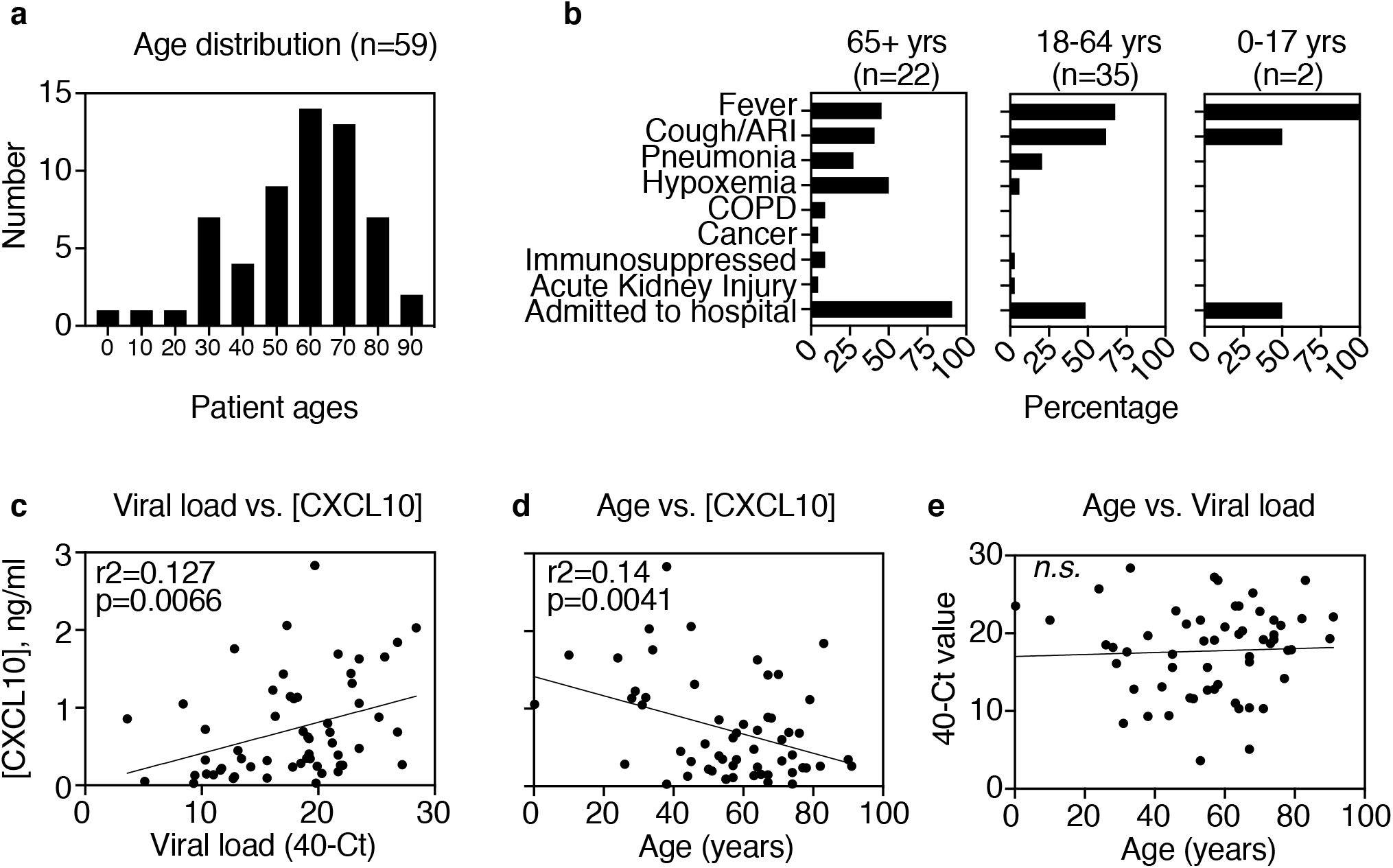
Clinical features and relationship to NP CXCL10 in SARS-CoV-2+ patients at YNHH (n=59) **(A)** Age distribution of all SARS-CoV-2+ patients in this study. **(B)** Clinical presentation by age group. Bar indicates percentage of patients in each age category displaying the clinical features listed on the y-axis. **(C)** Relationship between viral load, expressed at 40-Ct value for N1 gene, and NP [CXCL10] concentration. **(D)** Relationship between patient age and NP [CXCL10]. **(E)** Relationship between age and viral load. **(C-E)** Two-tailed Pearson correlation analysis was performed and r-squared values and p values are indicated for significant associations. No significant association was observed between age and viral load for these patients *(n.s.)*.

We also considered that NP CXCL10 level might indicate more robust antiviral responses in the nasal mucosa and therefore might correlate with biological variables associated with lower illness severity. Samples in this study were from patients of a wide age range (Fig 4a, b), enabling analysis of the relationship between viral load and age. Pearson correlation analysis showed a significant negative association between NP CXCL10 and age, with higher NP CXCL10 associated with younger age (Fig 4d). One explanation for the trend towards higher CXCL10 levels in younger patients might be that younger patients had higher NP viral loads; however, this did not appear to be the case as there was no clear correlation between age and viral RNA level in this sample set (Fig 4e). An alternate explanation could be that that the antiviral response in the upper respiratory tract is more robust in young patients compared to older patients during SARS-CoV-2 infection. It is also possible that high NP CXCL10 tracks with lower severity of illness; however, since all older patients in this sample set had more severe symptoms and were more likely to be hospitalized (Fig 4b) it was not possible to unlink age from illness severity in this analysis. Further work will be needed to examine this more fully.

Taken together, the results reported here indicate the potential for host-response based screening to help solve current challenges in diagnostic testing presented by the SARS-CoV-2 pandemic. One major challenge is the need to rapidly expand testing capacity to support surveillance as social distancing measures ease. We show that NP CXCL10 level had a very high negative predictive value for SARS-CoV-2 infection in patients with a range of symptoms, in which the COVID-19 prevalence was 12% (17/144) (Fig 3). This indicates that, if used as a pre-screen in a similar patient population, this test could potentially eliminate the need for ~80% of PCR tests by screening out samples very likely to be negative for the virus. Although this strategy may not capture every positive case, there also may be some cases in which biomarker based testing is more sensitive than PCR. Based on longitudinal studies, the sensitivity of PCR-based testing can vary considerably even on sequential days in the same patient^15-17^. It is possible that some of the 29 RVP-negative, CXCL10 positive patients in Figure 1 that tested negative for SARS-CoV-2 did indeed have COVID-19; however due to study design it is not possible to follow up with patients to further assess. Future longitudinal studies will enable better assessment of the relative sensitivity of CXCL10 and viral PCR for diagnosing COVID-19.

This study focused on symptomatic patients. In future studies, it will be important to assess biomarker performance in other populations, particularly if the intention is to screen populations who may have low NP viral loads (e.g. asymptomatic subjects.) Previous studies have shown that induction of host interferon stimulated genes occurs in the nasal mucosa of asymptomatic subjects with respiratory virus infection, but the level of induction may be lower^2,18-21^. For evaluating such subjects, it will be important to use a CXCL10 assay with a lower limit of detection than the one used here.

The link between viral replication level and biomarker expression suggests another potential use of biomarkers: assessing transmission risk. Recent work has highlighted persistent NP PCR positivity in patients up to 4 weeks post recovery from COVID-19^22,23^. This has raised questions about whether these positive tests are detecting infectious virus and a transmission risk, or are simply detecting persistent viral genome fragments or very low levels of virus which are not a transmission risk. A recent report from South Korea found that no transmission occurred to 790 contacts of 285 COVID-19 patients who tested positive for viral RNA post recovery^24^. Since the trigger for CXCL10 production is viral replication, it is possible that this biomarker could serve as a correlate of infectivity, which could be assessed using viral culture. Finding a biomarker to distinguish whether PCR-positivity signifies live/infectious virus will be particularly useful as very sensitive but non-quantitative tests for viral genetic material gain more widespread use, such as in-home point of care tests.

In conclusion, we provide evidence based on early cases of COVID-19 in our region that host biomarker-based screening has tremendous potential to solve some of the current challenges presented by the COVID-19 pandemic, including rapidly increasing testing capacity. While diagnostics that detect viral RNA are highly specific, these tests are complex, relatively expensive, and subject to supply chain interruption. In contrast, immunoassay can provide inexpensive, high throughput testing and can be easily adapted to point of care testing. While we previously identified several proteins highly induced during the antiviral interferon response, we focused on NP CXCL10 for this study as it is a well-known molecule for which validated detection antibodies and automated detection platforms already exist. While more study is needed, the work presented here demonstrates the great potential for biomarker-based screening to enable rapid expansion of testing capacity by directing existing testing to samples most likely to be virus-positive.

## Materials and Methods

### Human subjects

Residual nasopharyngeal samples from clinical testing were obtained from the Yale-New Haven Hospital Clinical Virology Laboratory and medical records were reviewed, data de-identified, and discovered positive cases were reported to health care providers according to IRB-approved protocol #2000027656 with oversight from the Yale Human Investigations Committee.

### Virology testing

For testing by the YNHH Clinical Virology Laboratory, NP swabs were placed in viral transport media (BD Universal Viral Transport Medium) immediately upon collection. Samples (200 μL) were subjected to total nucleic acid extraction using the NUCLISENS easyMAG platform (BioMérieux, France). The 15-virus PCR panel was performed as previously described, with updated rhinovirus PCR detection and inclusion of 4 seasonal coronaviruses and parainfluenza virus 4 ^2,25,26^ YNHH testing for SARS-CoV-2 was done using N1, N2, and RNAse P primer probe sets with an emergency use authorized assay based the CDC protocol.^27^

For screening of 642 respiratory virus panel negative samples for SARS-CoV-2 RNA, eluates from easyMag RNA extraction were screened using the US CDC 2019-nCoV N1 primer probe set or the E gene Sarbeco primer probe set, using the following reaction conditions as described previously^28,29^ (IDT, Coralville, Iowa). We used the Luna Universal Probe One-step RT-qPCR kit (New England Biolabs, Ipswich, MA, USA) with 5 μL of RNA and primer and probe concentrations of 500 nM of forward and reverse primer, and 250 nM of probe. PCR cycler conditions were reverse transcription for 10 minutes at 55°C, initial denaturation for 1 min at 95°C, followed by 40 cycles of 10 seconds at 95°C and 20 seconds at 55°C on the Biorad CFX96 qPCR machine (Biorad, Hercules, CA, USA). PCR-positive samples were confirmed by the YNHH clinical laboratories as described above.

### CXCL10 measurements

Human CXCL10 was measured in duplicate for each sample using the R&D Human CXCL10/IP-10 DuoSet ELISA (Cat No: DY266) and concentrations were calculated from a standard curve on each plate according to manufacturer instructions using GraphPad Prism software.

### Virus sequencing and phylogenetic analysis

SARS-CoV-2 positive samples were processed for next-generation sequencing as previously described^8^. Total nucleic acid was subjected to cDNA synthesis using SuperScript IV VILO Master Mix (ThermoFischer Scientific, MA, USA) according to the manufacturer’s protocol. cDNA was used as input into a highly multiplexed amplicon generation approach for sequencing on the Oxford Nanopore Technologies MinION (ONT, Oxford, UK)^30^. Samples were barcoded using the Native Barcoding Expansion Pack (ONT, Oxford, UK), multiplexed, and sequenced using R9.4.1 flow cells (ONT, Oxford, UK). The RAMPART software from the ARTIC Network was used to monitor each sequencing run. Runs were stopped when sufficient depth of coverage was achieved to accurately generate a consensus sequence. Following the completion of each sequencing run, raw reads (.fast5 files) were basecalled using Guppy high-accuracy model (v3.5.1, ONT, Oxford, UK). Basecalled FASTQ files were used as input into the ARTIC Networks consensus sequence generation bioinformatic pipeline (https://artic.network/ncov-2019/ncov2019-bioinformatics-sop.html). Variants to the reference genome were called with nanopolish^31^. Stretches of the genome that were not covered by 20 or more reads were represented by stretches of NNN’s.

To infer the evolutionary history and origins of the early sampled SARS-CoV-2 genomes, we performed phylogenetic analysis using a nextstrain pipeline^32^. Along its workflow, sequences were aligned using MAFFT^33^, and the phylogeny was inferred using a Maximum Likelihood approach implemented on IQTree^34^, with GTR substitution model. Ancestral continental origins were inferred as discrete characters using TreeTime^35^. Finally, the phylogenetic data visualization was obtained using Auspice^32^.

### Statistical analysis

Statistical analysis was performed using Pearson correlation analysis, twotailed, with Graph Pad Prism software.

## Data Availability

All data is provided in the manuscript. Additional details are available upon request.

## Acknowledgements

We would like to thank Maureen Owen, Robin Gardner, Greta Edelman, and the rest of the Yale-New Haven Hospital clinical virology laboratory for valuable assistance and support. We would like to thank Amelia Hanron, Timothy Watkins, Bao Wang, and Valia Mihaylova, for help and feedback. Funding was provided by the N.I.H. (K08 AI119139 to E.F.F.) the Hartwell Foundation (to E.F.F.), the Yale Department of Laboratory Medicine (to E.F.F.), the Yale School of Public health (to N.D.G), and the Huffman Family Donor Advised Fund (to N.D.G). CBFV is supported by NWO Rubicon 019.181EN.004.

## Author contributions

E.F. and N.C. conceived of the study. E.F., N.C., A.F.B, J.R.F., T.A., C.B.F.V., and N.D.G. designed and performed laboratory experiments and data analysis. E.F. and M.L. designed the clinical protocol. M.L. provided clinical samples and information and contributed to data interpretation. All authors gave scientific input. E.F. wrote the first draft of the manuscript. All authors reviewed and approved the final manuscript.

## Supplemental Figures

**Fig S1.**
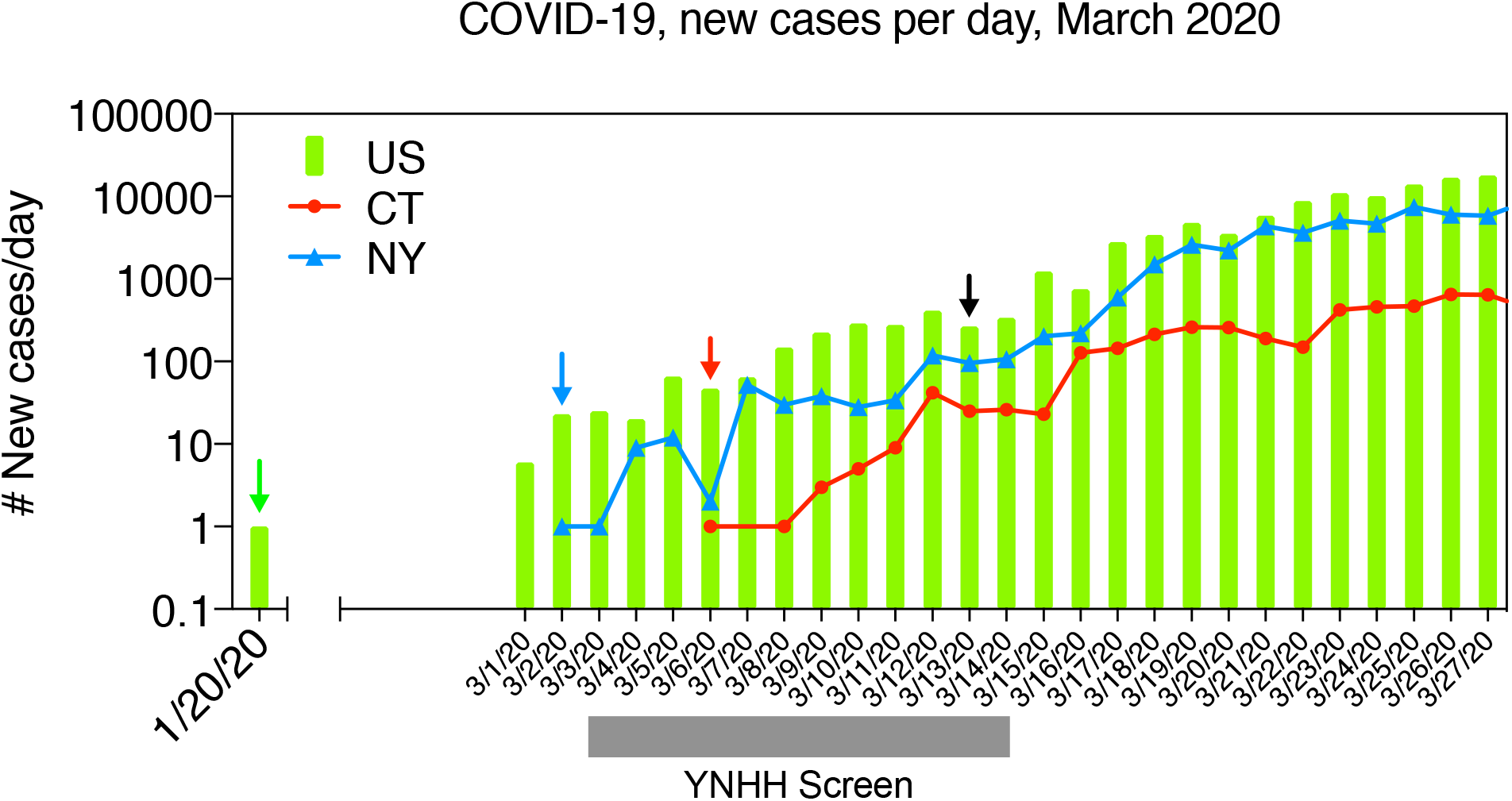
COVID-19 cases in U.S. in March 2020. New cases per day are shown for US (green bars), NY state (blue line) and CT (red line). Arrows indicate first reported case in U.S. (green), New York (blue), CT (red). Black arrow indicates start date of COVI19 testing at Yale-New Haven Hospital (YNHH). Grey bar indicates time window of screen for undiagnosed cases at YNHH shown in Figure 1.

**Table S1.** PCR tests on YNHH respiratory virus panel, 2020.

|  |
| --- |
| Rhinovirus<br>Coronaviruses (CoV-NL63, 229E, OC43, HKU1)<br>Influenza A and B<br>Parainfluenza 1, 2, 3, and 4<br>Respiratory syncytial virus A and B<br>Human metapneumovirus<br>Adenoviruses |

**Table S2.** Summary statistics for MinION sequencing of SARS-CoV-2 positive samples.

| Sample ID | Sample type | CT Value | Reads to Barcode | Reads to SARS-CoV-2 Genome | % Reads Aligned | Genome Coverage | Avg DOC<sup>b</sup> |
| --- | --- | --- | --- | --- | --- | --- | --- |
| Yale-009 | NP<sup>a</sup> | 21 | 123,547 | 123,543 | 100.0 | 97.4 | 200+ |
| Yale-011 | NP | 28 | 159,675 | 156,799 | 98.2 | 96.8 | 200+ |
| Yale-040 | NP | 22 | 345,561 | 342,796 | 99.2 | 98.6 | 200+ |
| Yale-151 | NP | 15 | 243,524 | 239,856 | 98.4 | 99.6 | 200+ |

a- NP= nasopharyngeal swab
b- DOC=Depth of coverage in reads across SARS-CoV-2 genome

**Table S3. List of genomes used in the phylogenetic analysis included in Fig 2**

Provided as a separate Excel file.

